# Hyperintense Acute Reperfusion Marker Sign in Patients with Diffusion Weighted Image-negative Transient Ischemic Attack HARM Sign in DWI-negative TIA

**DOI:** 10.1101/2024.06.16.24309010

**Authors:** Taewoo Kim, Bum Joon Kim, Ho Geol Woo, Kyung Mi Lee, Sangil Park, Dae-il Chang, Sung Hyuk Heo

## Abstract

**Background:** The hyperintense acute reperfusion marker (HARM) sign is a hyperintense signal observed on postcontrast fluid-attenuated recovery inversion images and is strongly associated with cerebral ischemic insults. The clinical significance of the HARM sign in transient ischemic attack (TIA) has rarely been studied, unlike that in stroke. This study investigated the association between the HARM sign and clinical factors of diffusion-weighted imaging (DWI)-negative TIA, and the relationship between the HARM sign and recurrence of TIA and ischemic stroke.

**Methods:** We included 329 consecutive patients with DWI-negative TIA and divided them into two groups according to the HARM sign: 299 patients in the HARM(-) group and 30 patients in the HARM(+) group. Clinical information, brain imaging and follow-up data were collected from medical records and phone calls, and compared using HARM sign.

**Results:** The HARM(+) patients were older and had higher systolic blood pressure, shorter symptom duration, and more frequent history of recent TIA or stroke and symptomatic artery stenosis or occlusion. Multivariate logistic regression revealed that recent TIA or stroke within 12 months (OR 6.623), symptom duration under 1 hour (OR 2.735), and relevant artery stenosis or occlusion (OR 2.761) were independently associated with the HARM sign. Cortical symptoms including aphasia were more prevalent in the HARM(+) group. During follow-up, HARM(+) patients showed higher recurrence rates of ischemic stroke (13.3% vs. 3.0%, p = 0.023). However, multivariate Cox analysis indicated that symptomatic stenosis or occlusion, rather than the HARM sign, was independently associated with stroke recurrence.

**Conclusion:** The HARM sign in DWI-negative TIA patients is linked to older age, recent cerebrovascular events, shorter symptom duration, and large artery stenosis or occlusion. While the HARM sign correlates with higher recurrence of ischemic stroke, large artery stenosis or occlusion is the primary independent predictor.

## Introduction

Transient ischemic attack (TIA) is a transient focal cerebral dysfunction of vascular origin, with a rapid onset and symptoms that typically last 2–15 minutes, but sometimes up to 1 day (24 hours).^1^ TIA is diagnosed based on transient neurological symptoms, and only 30% of patients with TIA are positive for such symptoms on brain diffusion-weighted imaging (DWI);^2^ thus, there is no definitive diagnostic tool for TIA.

Hyperintense acute reperfusion marker (HARM) is the contrast extravasation phenomenon, which denotes the radiologic finding of the hyperintense signal within the cerebrospinal fluid spaces visualized on a postcontrast fluid-attenuated inversion recovery (FLAIR) image, and indicates a blood-brain barrier (BBB) disruption.^3^ A study showed that, when more than 20,000 brain magnetic resonance imaging (MRI) scans were examined, the HARM sign was a rare finding identified in only 0.35% of the scans.^4^ However, the HARM sign was observed in 83% of cases of subacute strokes and 10% of TIA cases, indicating that the HARM sign can confirm a cerebral ischemic insult.^5,6^

In terms of clinical significance, the HARM sign correlates with hemorrhagic transformation and parenchymal hemorrhage, and it is associated with worse clinical outcomes of stroke.^7,8^ There also appears to be a close association between the HARM sign and stroke etiology in patients with acute ischemic stroke.^9^ Regarding TIA and the HARM sign, there was a small study, including 77 patients without diffusion abnormalities presenting with symptoms of acute stroke, in which the HARM sign was associated with older age and higher National Institute of Health Stroke Scale (NIHSS) score at admission and seemed to be associated with cortical symptoms.^10^ However, no significant follow-up studies have been conducted.

This study aimed to explore the association between HARM sign and clinical factors in patients with TIA, including demographic data, neurological symptoms, brain imaging and various risk factors. Furthermore, we investigated the relationship between HARM sign with recurrence of TIA and ischemic stroke through prospective follow-up and multivariate analysis.

## Materials and Methods

### 1. Patient selection and study design

This study included 520 consecutive patients admitted to Kyung Hee University Hospital between January 2015 and March 2023 diagnosed with TIA at discharge (Figure 1). Patients presenting with headache as the chief complaint and neurological symptoms lasting more than 24 hours (e.g., posterior reversible encephalopathy and reversible cerebral vasoconstriction syndrome) were excluded at the beginning of the study. We attempted to exclude TIA mimics as much as possible through a chart review at the time of admission and subsequent follow-up. Among 40 patients with the TIA mimics, 28 patients had transient global amnesia (TGA), with 2 DWI-positive TGAs and 26 DWI-negative TGAs. TGA cases were excluded from the study because previous studies have reported that the pathophysiological mechanisms of TGA and TIA are different.^11,12^ A total of 375 patients diagnosed with TIA were included in this study, of whom 329 were DWI-negative and 46 were DWI-positive. The 329 DWI-negative patients with TIA were divided into two groups based on the presence or absence of the HARM sign: 299 patients were in the HARM(-) group and 30 were in the HARM(+) group. The primary analysis in this study was a comparison between the two groups.

**Figure 1.**
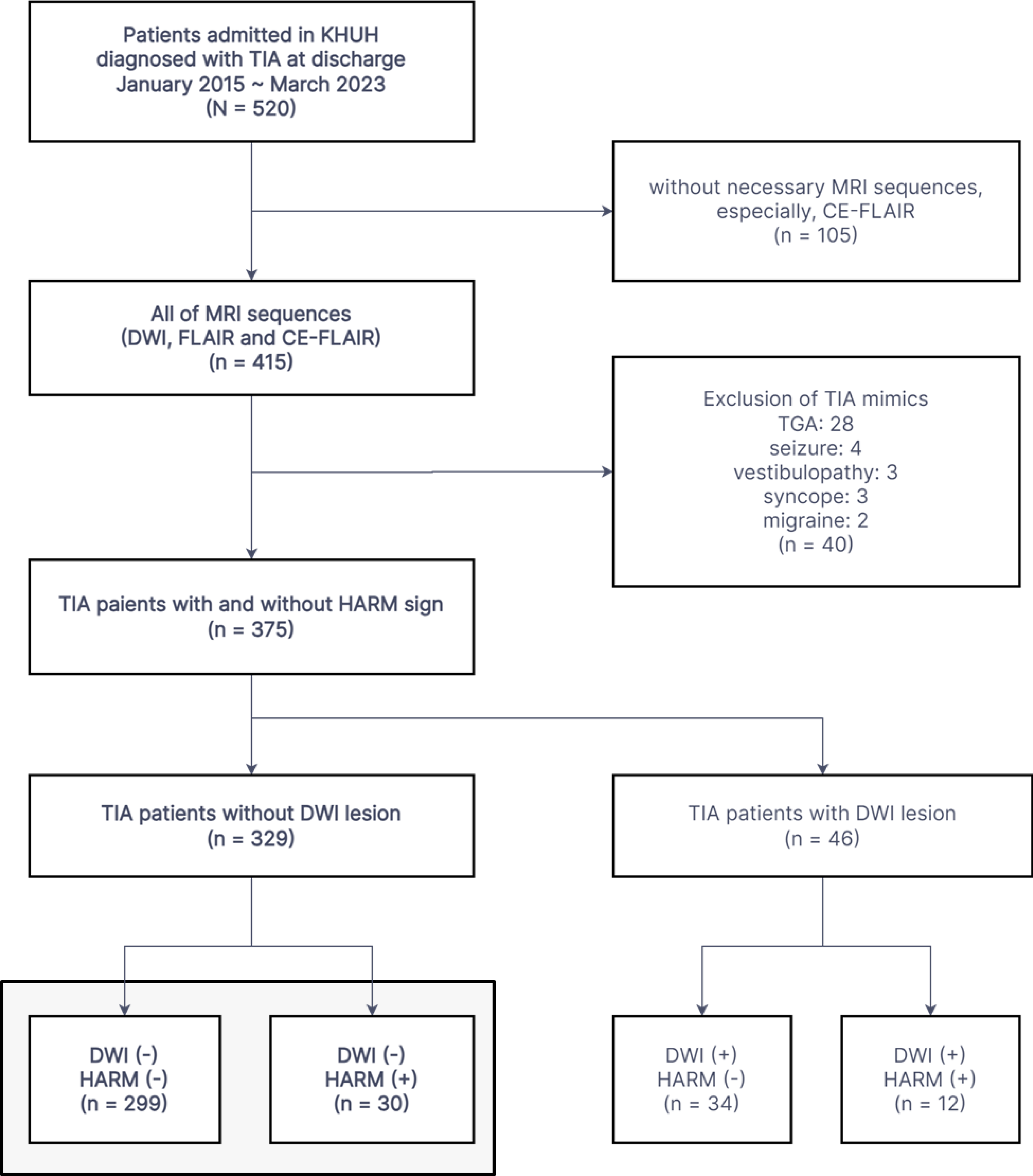
Flow chart of patient selection in this study. KHUH, Kyung Hee University Hospital; TIA, transient ischemic attack; DWI, diffusion-weighted imaging; FLAIR, fluid-attenuated inversion recovery imaging; CE-FLAIR, contrast-enhanced FLAIR; HARM, hyperintense acute reperfusion marker on CE-FLAIR; TGA, transient global amnesia.

This study was approved by the Independent Ethics Committee of Kyung Hee University Medical Center (KMC IRB 2023-11-053).

### 2. Clinical information

All clinical information was obtained from the medical records of Kyung Hee University Hospital. Data included demographic characteristics, medical history, medications, laboratory tests, ABCD2 scores, National NIHSS scores, and stroke workup.

Based on the medical records of the patients, the symptoms were categorized as follows: cortical symptoms (aphasia, neglect, confusion, and homonymous hemianopsia), lateralized motor weakness, lateralized sensory symptoms, dizziness/vertigo, diplopia, transient monocular blindness, and non-localized symptoms (dysarthria, non-lateralized motor symptoms, and loss of consciousness). Patients may experience multiple symptoms. Because of the nature of TIA, patients often improved before they came to the hospital; therefore, clinicians frequently relied on the statements of the patients and caregivers and not an examination to collect clinical information.

### 3. Image acquisition and analysis

MRI was performed using a 3 Tesla MRI system (VIDA, Siemens or Achieva TX; Philips Medical Systems, Best, The Netherlands) equipped with a 64- or 32-channel head coil. The protocol for conventional brain MRI evaluation included DWI, ADC maps, three-dimensional magnetization-prepared rapid gradient echo T1WI (3D MPRAGE T1WI), T2WI, FLAIR, susceptibility-weighted imaging, 3D postcontrast T1WI, postcontrast FLAIR, and vascular imaging, including time-of-flight intracranial MRA and postcontrast MRA encompassing the extracranial internal carotid and vertebral arteries. For enhancing images, patients were administered gadolinium (Prohance, Bracco Diagnostics, Princeton, NJ, USA) at a dose of 0.2 mmol/kg, and postcontrast FLAIR images were obtained 5–8 min later.

Brain MRI images were evaluated by two experienced stroke neurologists to determine DWI positivity and the presence of the HARM sign. They corroborated the lesion identification of the HARM on abnormal cortical and sulcal enhancements of FLAIR images by reviewing other sequence images. Disagreements between the two neurologists were resolved by one neuroradiologist.

All patients with TIA underwent brain MRA. The major intracranial and extracranial arteries were assessed for stenosis and occlusion. Based on major studies including patients with TIA, such as the WASID and OXVASC studies,^13,14^ symptomatic stenosis of 50–99% was the target of this study and was subclassified into severe stenosis (70–99%). Stenosis was assessed using the WASID method (between the narrowest point and compared with the luminal size before the stenosis).^15^ Symptomatic stenosis and occlusion were demonstrated to correlate with clinical presentation and brain imaging, as in other studies.^13,14,16^ Extracranial arteries assessed included subclavian, common carotid, proximal internal carotid, and vertebral arteries (V1, V2, and V3), and intracranial arteries assessed included distal internal carotid, middle cerebral (M1 and M2), anterior cerebral (A1), posterior cerebral (P1), and basilar and vertebral arteries (V4). These were also evaluated by two-stroke neurologists, and in cases of disagreement, one neuroradiologist confirmed the diagnosis.

### 4. Follow-up data

Patients with TIA were prospectively followed from discharge from the hospital until August 2023. For patients being followed up during the outpatient visit, we referred to the medical records and imaging examinations of the hospital and obtained clinical information regarding the recurrence of TIA or stroke, administration of antithrombotic agents, and follow-up duration. Patients lost to follow-up were contacted by phone and asked about recurrent symptoms, newly diagnosed TIA or stroke, and administration of antithrombotic agents. We also evaluated all patients with TIA for a new diagnosis of atrial fibrillation after discharge from the hospital.

### 5. Statistical analysis

For continuous variables, results are presented as mean ± standard deviation or median (interquartile range), whereas for categorical variables, results are expressed as the number of participants (%). Chi-square and Fisher’s exact tests were used to compare categorical variables. The independent t-test was used for parametric analysis, and the Mann–Whitney U test was used for non-parametric analysis. To evaluate independent risk factors for the HARM sign, variables of P < 0.1 from univariate logistic regression analysis were input to multivariate logistic regression analysis using the “enter” method. Odds ratios (ORs) and 95% confidence intervals (CIs) were also determined. The cumulative incidences of recurrent TIA and stroke were estimated using the Kaplan-Meier method, and the curves were compared using the log-rank test. Risk ratios for stroke recurrence were estimated as hazard ratios using the Cox proportional hazards model. Major variables commonly associated with recurrent stroke were analyzed using univariate Cox analysis, and multivariate Cox analysis was performed for variables with p < 0.1 in the univariate analysis. The hazard ratio and 95% CI were obtained. Multicollinearity and potential interactions among variables were examined by assessing the variance inflation factor. SPSS software was used for the analysis (SPSS version 27.0, IBM Corp., Chicago, IL, USA), and a p-value of <.05 was set for statistical significance.

## Results

### 1. Baseline characteristics of study participants

We identified 375 consecutive patients with TIA, 329 of whom were DWI negative, as previously described (Figure 1). DWI lesions were found in 46 patients (12.3%), and 42 patients (11.2%; 12 with DWI lesions and 30 without) had HARM signs corresponding to neurological symptoms (Figure S1). The baseline characteristics of the DWI-negative TIA group were compared and summarized according to the presence of the HARM sign (Table 1). The HARM(+) group was older than the HARM(-) group. A history of previous cerebral insults, including TIA and stroke, was more common in the HARM(+) group (43.3% vs. 22.1%, p = 0.009). Especially when limited to TIA and stroke within 12 months, the difference between the groups was larger and more statistically significant (26.7% vs. 4.0%, p < 0.001).

**Table 1.** Baseline characteristics of patients with DWI-negative TIA and their comparisons by HARM sign.

| Variables | HARM negative<br>(n = 299) | HARM positive<br>(n = 30) | p-value |
| --- | --- | --- | --- |
| Age | **64.4 ± 11.9** | **70.7 ± 13.5** | **0.007** |
| Male sex | 150 (50.2%) | 16 (53.3%) | 0.741 |
| Medical history |  |  |  |
| Cerebral insult | **66 (22.1%)** | **13 (43.3%)** | **0.009** |
| TIA | 26 (8.7%) | 5 (16.7%) | 0.154 |
| Stroke* | **48 (16.1%)** | **10 (33.3%)** | **0.018** |
| Previous TIA or stroke history<br>within 12 months | **12 (4.0%)** | **8 (26.7%)** | **<0.001** |
| Hypertension | 184 (61.5%) | 23 (76.7%) | 0.102 |
| Diabetes | 79 (26.4%) | 8 (26.7%) | 0.977 |
| Dyslipidemia | 184 (61.5%) | 16 (53.3%) | 0.380 |
| Smoking (current or quit < 5 years) | 62 (20.7%) | 7 (23.3%) | 0.739 |
| Atrial fibrillation | 27 (9.0%) | 4 (13.3%) | 0.442 |
| Taking antiplatelet at the onset | 94 (31.4%) | 11 (36.7%) | 0.558 |
| Taking anticoagulant at the onset | 11 (3.7%) | 3 (10.0%) | 0.102 |
| Body mass index | 24.55±3.15 | 25.07±4.98 | 0.584 |
| Laboratory test |  |  |  |
| WBC | 7.06 ± 2.36 | 7.59 ± 4.06 | 0.282 |
| BUN | 17.10 ± 6.45 | 19.47 ± 6.75 | 0.057 |
| Creatinine | **0.84 ± 0.29** | **0.97 ± 0.31** | **0.018** |
| Hemoglobin | 13.72 ± 1.60 | 13.35 ± 1.73 | 0.234 |
| Hematocrit | 40.3 ± 4.3 | 39.4 ± 5.1 | 0.260 |
| Triglyceride | 147.5 ± 110.2 | 140.1 ± 87.4 | 0.725 |
| Total cholesterol | 175.7 ± 44.0 | 182.3 ± 54.2 | 0.446 |
| HDL cholesterol | 48.7 ± 13.8 | 51.2 ± 14.7 | 0.344 |
| LDL cholesterol | 104.7 ± 34.8 | 109.7 ± 39.0 | 0.463 |
| HbA1c | 6.02 ± 1.00 | 6.15 ± 1.35 | 0.522 |
| Initial systolic BP | **144.1 ± 22.6** | **154.3 ± 27.0** | **0.022** |
| Initial diastolic BP | 82.3 ± 13.3 | 82.5 ± 12.6 | 0.928 |
| ABCD2 score | 4 [3, 5] | 4 [3, 6] | 0.991 |
| Duration of symptoms under 1 h | **116 (38.8%)** | **19 (63.3%)** | **0.009** |
| Baseline NIHSS score | 0 [0, 2] | 0 [0, 1] | 0.255 |
| Discharge NIHSS score | 0 [0, 0] | 0 [0, 0] | 0.425 |
| Stroke workup |  |  |  |
| MR angiography | 299 (100%) | 30 (100%) |  |
| Symptomatic stenosis (50–99%) or occlusion | **42 (14.0%)** | **18 (60.0%)** | **<0.001** |
| Symptomatic stenosis (50–99%) | **28 (9.4%)** | **11 (36.7%)** | **<0.001** |
| Occlusion | **14 (4.7%)** | **7 (23.3%)** | **0.001** |
| Routine ECG | 299 (100%) | 30 (100%) |  |
| 24-hour Holter monitoring | 88 (29.4%) | 13 (43.3%) | 0.116 |
| TTE | 197 (65.9%) | 23 (76.7%) | 0.232 |

TIA, transient ischemic attack; WBC, white blood cell; BUN, blood urea nitrogen; NIHSS, National Institute of Health Stroke Scale; HARM, hyperintense acute reperfusion marker; BP, blood pressure; TTE, transthoracic echocardiogram
p-values were estimated using the Pearson chi-square test, Fisher's exact test, Mann–Whitney U test, or independent t-test, as appropriate.
\* Stroke included cerebral infarction, spontaneous intracerebral hemorrhage, and traumatic intracranial hemorrhage.

Initial systolic blood pressure (BP) were higher in the HARM(+) group than in the HARM(-) group. Patients with a symptom duration of less than 1 hour was higher in the HARM(+) group (63.3% vs. 38.8%, p = 0.009), and symptomatic stenosis (50-99%) or occlusion was much more prevalent in the HARM(+) group (60.0% vs. 14.0%, p < 0.001). The proportions of moderate and severe stenosis and occlusion in the steno-occlusive lesions were not significantly different between the two groups. There were no statistically significant differences in other baseline characteristics, including ABCD2 and NIHSSscores, between the two groups.

Logistic regression analysis was performed on the seven variables with p < 0.1 shown in Table 1 (Table 2). In the univariate analysis, the p-values of the variables were similar to those shown in Table 1. In multivariate logistic regression analysis, previous TIA or stroke histories within 12 months (OR 6.623, 95% CI, 2.206–19.880), duration of symptoms under 1 hour (OR 2.735, 95% CI, 1.148–6.512), and relevant artery stenosis (≥50%) or occlusion (OR 761, 95% CI, 1.159–6.580) were independently associated with HARM sign in DWI-negative TIA cases.

**Table 2.** Logistic regression analysis for association between HARM sign and baseline characteristics of patients with TIA.

| Variables | Univariate |  | Multivariate |  |
| --- | --- | --- | --- | --- |
|  | Odds ratio (95% CI) | p-value | Odds ratio (95% CI) | p-value |
| Age | 1.048 (1.013-1.085) | 0.008 | 1.029 (0.989–1.071) | 0.157 |
| Previous TIA or stroke history within 12 months | 8.697 (3.218-23.503) | <0.001 | **6.623 (2.206–19.880)** | **<0.001** |
| BUN | 1.047 (0.997-1.098) | 0.063 | 0.998 (0.927–1.074) | 0.951 |
| Creatinine | 3.290 (1.181-9.164) | 0.023 | 1.772 (0.399–7.874) | 0.452 |
| Initial systolic BP | 1.018 (1.002-1.033) | 0.024 | 1.014 (0.998–1.031) | 0.083 |
| Duration of symptoms under 1 hour | 2.725 (1.251-5.933) | 0.012 | **2.735 (1.148–6.512)** | **0.023** |
| Symptomatic stenosis (50–99%) or occlusion | 4.691 (2.146-10.254) | <0.001 | **2.761 (1.159–6.580)** | **0.022** |

The major variables in Table 1 (p-values less than 0.1) were selected for univariate and multivariate analyses.

### 2. Clinical symptoms in patients with TIA

We categorized TIA symptoms and compared their association with HARM sign (Table 3). Cortical symptoms, such as aphasia, neglect, confusion, and homonymous hemianopsia, were more prevalent in the HARM(+) group than in the HARM(-) group; this difference was statistically significant (30.0% vs. 8.7%, p = 0.002). Other symptoms did not significantly differ between the two groups. Non-localized symptoms such as dysarthria, non-lateralized motor symptoms, and loss of consciousness were reported in 106 patients (8 with HARM and 98 without HARM). Of these, 36 patients had isolated non-localized symptoms, meaning no additional symptoms, such as lateralized weakness or cortical symptoms (2 with HARM and 34 without HARM).

**Table 3.** Transient neurological symptoms of patients with DWI-negative TIA and their comparisons by HARM sign.

| Clinical symptoms | HARM negative<br>(n = 299) | HARM positive<br>(n = 30) | p-value |
| --- | --- | --- | --- |
| Cortical symptoms | **26 (8.7%)** | **9 (30.0%)** | **0.002** |
| Aphasia | **20 (6.7%)** | **6 (20.0%)** | **0.005** |
| Neglect | 1 (0.3%) | 0 (0.0%) | >0.999 |
| Confusion | 7 (2.3%) | 3 (10.0%) | 0.053 |
| Homonymous hemianopsia | 2 (0.7%) | 0 (0.0%) | >0.999 |
| Lateralized motor weakness | 204 (68.2%) | 20 (66.7%) | 0.861 |
| Lateralized sensory symptoms | 69 (23.1%) | 4 (13.3%) | 0.221 |
| Dizziness/vertigo | 39 (13.0%) | 2 (6.7%) | 0.559 |
| Diplopia | 10 (3.3%) | 1 (3.3%) | >0.999 |
| Transient monocular blindness | 1 (0.3%) | 1 (3.3%) | 0.174 |
| Non-localized symptoms | 98 (32.8%) | 8 (26.7%) | 0.495 |
| Dysarthria | 93 (31.1%) | 8 (26.7%) | 0.615 |
| Non-lateralized motor symptom | 6 (2.0%) | 0 (0.0%) | >0.999 |
| Loss of consciousness | 3 (1.0%) | 0 (0.0%) | >0.999 |

P-values were estimated using Pearson's chi-square test or Fisher's exact test, as appropriate.

### 3. Recurrence of TIA and stroke in patients with TIA

A total of 329 patients with DWI-negative TIA were followed up from discharge to August 2023, and the two groups were compared based on the HARM sign (Table 4). The median total follow-up period was similar for both groups. The number of recurrent TIA and ischemic stroke occurrences during follow-up was higher in the HARM(+) group, but not statistically significant (16.7% vs. 7.0%, p = 0.074). When limited to ischemic stroke, the difference between the two groups was statistically significant (13.3% vs. 3.0%, p = 0.023). The number of patients taking antithrombotic agents was similar between the two groups.

**Table 4.** Follow-up data of patients with DWI-negative TIA and their comparisons by HARM sign.

| Variable | HARM negative<br>(n = 299) | HARM positive<br>(n = 30) | p-value |
| --- | --- | --- | --- |
| Total follow-up duration (months) | 37.0 [13.0, 65.0] | 36.0 [21.0, 59.0] | 0.781 |
| Follow-up loss within 3 months | 25 (8.4%) | 2 (6.7%) | 0.747 |
| 3 months mRS | 0 [0, 0] | 0 [0, 1] | 0.154 |
| Recurrent TIA or ischemic stroke | 21 (7.0%) | 5 (16.7%) | 0.074 |
| Recurrent ischemic stroke | **9 (3.0%)** | **4 (13.3%)** | **0.023** |
| Taking antithrombotic agent* | 233 (77.9%) | 25 (83.3%) | 0.493 |
| antiplatelet | 209 (69.9%) | 23 (76.7%) | 0.438 |
| anticoagulant | 30 (10.0%) | 4 (13.3%) | 0.571 |
| Atrial fibrillation confirmed until the last follow-up | 34 (11.4%) | 5 (16.7%) | 0.392 |

P-values were estimated using the Mann–Whitney U test, Pearson chi-square test, or Fisher’s exact test, as appropriate.
\* Taking an antithrombotic agent indicates whether the patient took medication at the time of their first recurrent TIA or stroke or the last follow-up.

The cumulative TIA and ischemic stroke recurrence was plotted using the Kaplan-Meier method and stratified by the HARM sign (Figure 2). The HARM(+) group had a higher TIA and ischemic stroke recurrence rate; however, this difference was not statistically significant (Figure 2A; log-rank test, p = 0.065). The cumulative recurrence of ischemic stroke was also analyzed using the Kaplan-Meier method, and the HARM (+) group had a significantly higher stroke recurrence rate (Figure 2B; log-rank test, p = 0.007).

**Figure 2.**
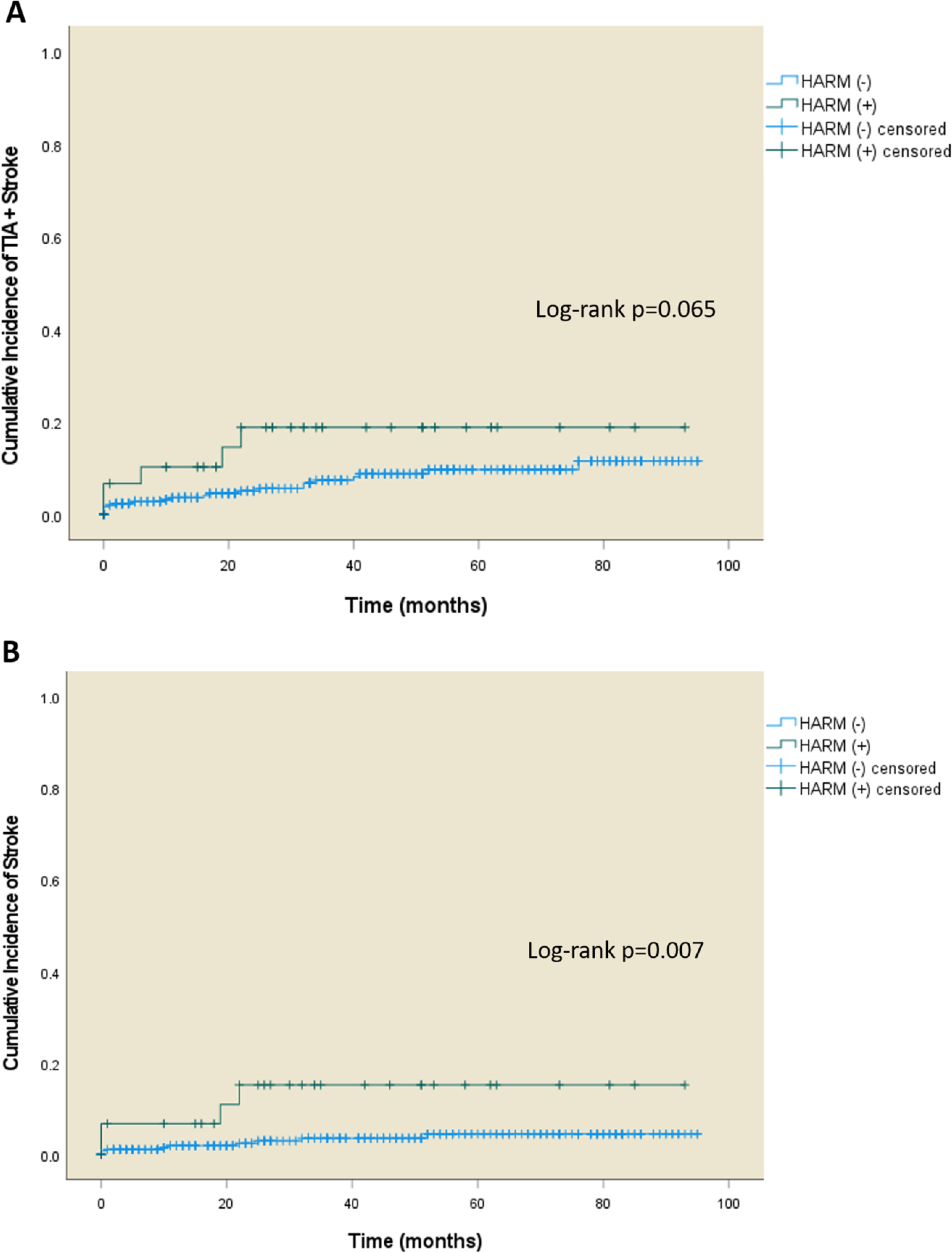
Kaplan-Meier analyses of cumulative incidence of (A) recurrent TIA or stroke, and (B) only recurrent stroke within patients with DWI-negative TIA with and without HARM sign. Log-rank tests were performed for each curve.

The Cox proportional model was used to analyze the independent variables associated with recurrent stroke in the DWI-negative TIA group (Table S1). Univariate analysis was performed with variables commonly associated with recurrent stroke, including the HARM sign, whose association with recurrent stroke in TIA was demonstrated previously using the log-rank test (Figure 2B). Multivariate Cox analysis was performed on four variables with p < 0.1 (age, ABCD2 score, relevant artery stenosis, and HARM sign) in the univariate analysis, and only symptomatic stenosis or occlusion was independently associated with recurrent stroke in DWI-negative TIA. Unlike in the univariate analysis, the HARM sign did not demonstrate an independent association with recurrent stroke in the multivariate analysis.

## Discussion

In this study, we retrospectively investigated the relationship between the HARM sign and various clinical factors in patients with DWI-negative TIA, which has not been explored before. Our study also investigates the association between the HARM sign and recurrent ischemic events of DWI-negative TIA prospectively. HARM sign was associated with older age, history of cerebral insult, initial high BP, and shorter duration of neurological symptoms in patients with TIA. In particular, recent TIA and stroke within 12 months, symptom duration within 1 hour, and large artery stenosis or occlusion were independently associated with HARM sign. Among the transient neurological symptoms in patients with TIA, cortical symptoms including aphasia seemed to be most closely associated with the HARM sign. Recurrent stroke in patients with TIA was correlated with the HARM sign in the univariate analysis; however, in the multivariate analysis, the HARM sign did not demonstrate statistical significance, whereas large artery stenosis and occlusion were independently associated with recurrent stroke.

Some studies have suggested that aging may be associated with BBB disruption, which is linked to cognitive decline in older adults.^17,18^ Another study also found that the BBB becomes more permeable with normal aging.^19^ Since the HARM sign is indicative of BBB disruption,^20^ it is possible that the HARM sign is more prevalent in older age groups. The older age of the HARM(+) group in this study is consistent with this reasoning. In addition, the DWI-negative patients with HARM sign in the previous study were older than those without HARM sign (73.1 vs. 55.9, p < 0.001),^10^ which is consistent with our results.

It has been reported in several studies that stroke causes BBB disruption.^5,21^ After ischemia, the activation of microglia and astrocytes leads to elevated levels of cytokines, chemokines, matrix metalloproteinases, and vascular endothelial growth factor within the ischemic brain tissue.^22^ These inflammatory agents and proteases are responsible for stroke-induced neuroinflammation and the apoptotic process of endothelial cells causing BBB breakdown.^21,22^ As in our study, recent TIAs and strokes could increase the vulnerability to BBB. If this is followed by subsequent cerebral ischemia, such as TIA, it may produce the HARM sign. However, it is possible that the inflammatory agents and proteases activated by ischemia remain upregulated for a prolonged period, thus making BBB breakdown more easily triggered by subsequent ischemia. Many of the agents involved in BBB disruption are known to increase over days to weeks.^22^ Furthermore, the fact that HARM signs are observed in 25% of patients with acute ischemic stroke,^9^ compared with 83% with subacute stroke,^5^ indicates that many factors that cause BBB breakdown may be upregulated for some time.

The correlation between symptom duration and the presence of the HARM sign was interesting. Intuitively, a longer symptom duration would suggest a longer duration of tissue ischemia and, therefore, increased BBB breakdown. However, this study found an independent association between a short symptom duration and HARM sign. In acute ischemic stroke, reperfusion is essential for the survival of brain tissue but paradoxically causes further damage to brain tissue including BBB disruption.^23,24^ Cerebral ischemia is also a risk factor for BBB disruption; however, reperfusion is the strongest independent predictor of early BBB disruption.^23^ Similarly, an animal study comparing rats that occluded the middle cerebral artery for 6 hours to another group that reperfused for 3 hours after 3 hours of occlusion found significantly more BBB disruption in the reperfusion group (p < 0.05).^25^ In several clinical practice studies, increased HARM sign after IV thrombolysis or mechanical thrombectomy has been found.^7,26^ This association between acute reperfusion therapy and HARM sign suggests that early robust reperfusion, meaning short duration of symptoms in this study, may cause more BBB disruption. The association of the HARM sign with high initial systolic BP in the univariate analysis in this study also suggests that robust reperfusion may be associated with BBB disruption.

Few previous studies on symptom duration in TIA have divided TIA into short- and long-duration TIA based on 1 hour, similar to our study.^27,28^ In a study by Weimer et al. including 1429 TIA patients, short-duration TIA had a significantly higher frequency of large artery stenosis, and the authors suggested that short- and long-duration TIA may have different pathogenesis.^28^ In this study, the HARM sign was closely associated with a short duration of symptoms and large artery stenosis or occlusion (Table 1). In addition, a further chi-square test for duration of symptoms under 1 hour and symptomatic stenosis or occlusion in our study showed a significant correlation (p = 0.015), which is consistent with the findings of Weimer et al.^28^ Further studies are needed to explain the significant correlation between HARM sign and short duration of symptoms in patients with TIA.

Among the main etiologies of stroke, large artery atherosclerosis and cardiogenic embolism were highly associated with the HARM sign (p < 0.001), while small vessel occlusion (SVO) was correlated with the absence of the HARM sign (p < 0.001) in a previous study.^9^ In our study, we found a causal relationship between large artery stenosis and the HARM sign, which is somewhat expected. The association of cortical symptoms with the HARM sign in our results is also consistent with this because cortical symptoms of ischemic stroke are themselves indicative of large artery involvement.^29^ It is also possible that subcortical lesions represented by SVO are anatomically unlikely to produce HARM sign. The HARM sign is thought to be observed when the contrast agent leaks from vessels due to BBB disruption, passes through the pia mater adjacent to the cortex and is distributed in the subarachnoid space.^30,31^ Therefore, we can speculate that the HARM(-) group is associated with SVO, and the HARM(+) group is associated with large artery atherosclerosis in our study.

However, the failure to demonstrate a relationship between the HARM sign and atrial fibrillation was unexpected. While 24-hour Holter monitoring is routinely performed when a patient with stroke is hospitalized, patients with TIA in this study were undertested, with 43% in the HARM(+) group and 29% in the HARM(-) group. It is possible that the diagnosis of paroxysmal atrial fibrillation was missed. Perhaps because of the asymptomatic nature of TIA, patients were unwilling to stay in the hospital for long periods and continue testing. In addition, although the sample size was small, the previous study did not demonstrate a significant association between the HARM sign and atrial fibrillation (p = 0.635), which is consistent with our findings.^10^ These findings were contrary to the previous study about the significant relationship between cardiogenic embolism and the HARM sign in ischemic stroke.^9^

HARM sign was associated with recurrent stroke using the univariate analysis, but did not demonstrate an independent relationship in the multivariate analysis. In this study, symptomatic stenosis and occlusion of the cerebral arteries were independently associated with recurrent stroke. This is consistent with previous studies in which large artery atherosclerosis was the etiology most associated with recurrent stroke.^32,33^ Since there were more patients with symptomatic stenosis and occlusion in the HARM (+) group in this study, it is thought that there were more recurrent strokes in the HARM(+) group. If cerebral atherosclerosis is present in patients with TIA and the HARM sign, more intensive treatment is required.

This study has some limitations. First, it used a small sample size and a non-randomized design in a single center. Therefore, the results are vulnerable to selection bias, provide restricted testing power, and limit generalizability. Second, as mentioned earlier, patients with TIA in this study did not undergo sufficient cardiac workups, including 24-hour Holter monitoring. Because nearly two-thirds of the patients did not undergo 24-hour Holter monitoring, the diagnosis of paroxysmal atrial fibrillation may have been missed. Future studies should include more participants and perform more thorough cardiac workups. Finally, most patients with DWI-negative TIAs were free of residual symptoms. In other words, it may be difficult to identify the symptomatic cerebral arteries. However, as mentioned earlier, only 36 patients (10.9%) had isolated non-localized symptoms, and most patients with TIA had additional symptoms with a localizing value. In addition, only four patients in the HARM(-) group had both isolated non-localized symptoms and major vessel stenosis; therefore, it was not a problem to identify symptomatic stenosis. It may be difficult to recognize cortical symptoms such as neglect or apraxia if these symptoms improve before the patient arrives at the hospital. Therefore, of the cortical symptoms presented in Table 3, various symptoms were usually identified using the statements of the patient or caregiver and could have been categorized just as “confusion.”

## Data Availability

The authors confirm that the data supporting the findings of this study are available within the article.

---

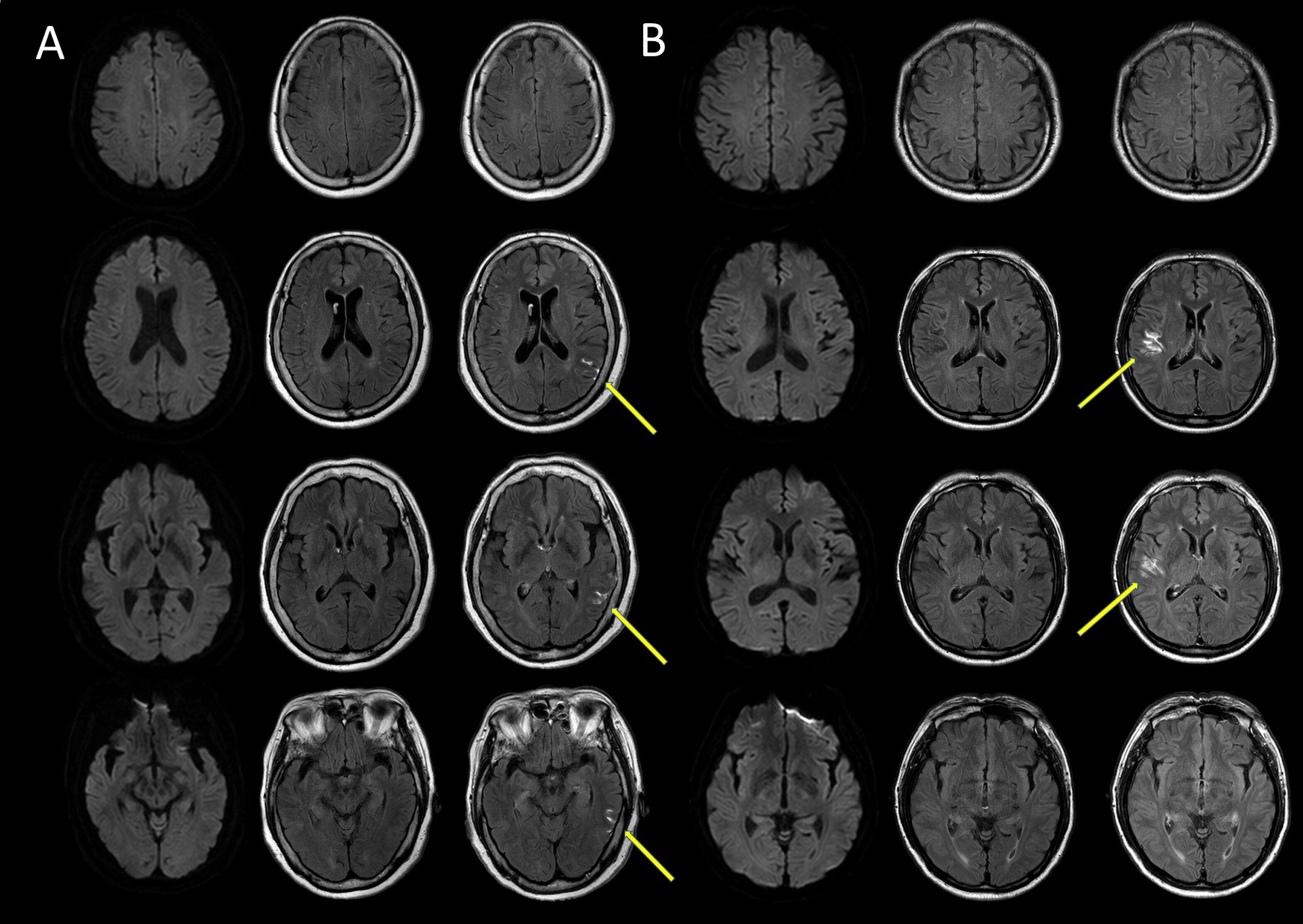

